# Bystander intervention is associated with reduced mortality among injury victims in Cameroon

**DOI:** 10.1101/2024.01.16.24301355

**Authors:** Kathleen O’Connor, Matthew Driban, Rasheedat Oke, Fanny Nadia Dissak-Delon, Signe Mary Magdalene Tanjong, Tchekep Mirene, Mbeya Dieudonne, Thompson Kinge, Richard L. Njock, Daniel N. Nkusu, Jean-Gustave Tsiagadigui, Cyrille Edouka, Catherine Wonja, Zachary Eisner, Peter Delaney, Catherine Julliard, Alain Chichom-Mefire, S. Ariane Christie

## Abstract

**Introduction:** Despite high injury mortality rates, Cameroon currently lacks a formal prehospital care system. In other sub-Saharan African low and middle-income countries, Lay First Responder (LFR) programs have trained non-medical professionals with high work-related exposure to injury in principles of basic trauma care. To develop a context-appropriate LFR program in Cameroon, we used trauma registry data to understand current layperson bystander involvement in prehospital care and explore associations between current non-formally trained bystander-provided prehospital care and clinical outcomes.

**Methods:** The Cameroon Trauma Registry (CTR) is a longitudinal, prospective, multisite trauma registry cohort capturing data on injured patients presenting to four hospitals in Cameroon. We assessed prevalence and patterns of prehospital scene care among all patients enrolled in a prospective, multisite trauma registry (Cameroon Trauma Registry (CTR)) in 2020. Associations between scene care, clinical status at presentation, and outcomes were tested using univariate and multivariate logistic regression. Injury severity was measured using the abbreviated injury score. Data were analyzed using Stata17.

**Results:** Of 2212 injured patients, 455 (21%) received prehospital care (PC) and 1699 (77%) did not receive care (NPC). Over 90% (424) of prehospital care was provided by persons without formal medical training. The most common prehospital interventions performed included bleeding control (370, 57%) and fracture immobilization (139, 21%). PC patients were more severely injured (p<0.001), had markers of increased socioeconomic status (p=0.01), and longer transport distances (p<0.001) compared to NPC patients. Despite increased severity of injury, patients who received PC were more likely to present with a palpable pulse (OR=6.2, p=0.02). Multivariate logistic regression adjusted for injury severity, socioeconomic status and travel distance found PC to be associated with reduced emergency department mortality (OR=0.14, p<0.0001).

**Conclusions:** Although prehospital injury care in Cameroon is rarely performed and is provided almost entirely by persons without formal medical training, prehospital intervention is associated with increased early survival after injury. Implementation of LFR training to strengthen the frequency and quality of prehospital care has considerable potential to improve trauma survival. LFR training in Cameroon should target commercial drivers given high work-related exposure to the injured and emphasize bleeding control interventions.

## INTRODUCTION

Low- and middle-income countries (LMICs) bear 90% of the world’s mortality burden due to injury. ^1^ Prehospital care is a critical component of the survival chain following injury, but prehospital care availability is extremely limited in LMICs. Trauma system development and quality improvement remain ongoing priorities in LMICs, with interest in prehospital care systems increasing as evidence emerges that basic prehospital life support measures may be equally effective as advanced interventions. ^2^ In May 2023, the 76th World Health Assembly (WHA) resolved that emergency, critical and operative care services are an integral part of a comprehensive national primary health care approach and foundational for health systems to effectively address emergencies. ^3^ Trauma system implementation may potentially address 54% of all-cause mortality across LMICs with integrated prehospital and inpatient emergency care. ^2,4,5^

The World Health Organization (WHO) affirms that prehospital care is a crucial component of mature trauma systems. In settings without existing prehospital infrastructure, the WHO has recommended training bystanders as Lay First Responders (LFRs) as the first step toward prehospital infrastructure development. ^6^ LFR programs have since emerged to train non-medical professionals with high work-related exposure to injury in basic principles of trauma prehospital care. Such trainee populations have included mototaxi drivers, taxi drivers, police officers, and firefighters. ^7-9^ Implementation of LFR programs may reduce 45% of all-cause mortality across LMIC. ^4^ Among LFR programs piloted across multiple sub-Saharan African countries, evaluations have been restricted to LFR educational outcomes, program cost effectiveness, and prehospital patient analyses without longitudinal clinical outcomes due to the lack of prospective data following LFR implementation. ^8, 10-12^ Without longitudinal clinical data to measure patient outcomes following prehospital LFR care, evaluation of LFR program efficacy has been limited.

In 2015, we developed the Cameroon Trauma Registry (CTR), a prospective, ongoing, multisite trauma registry that has since collected data on over 30,000 patients across four medical centers: Laquintinie Hospital, Regional Hospital of Limbe, Catholic Hospital of Pouma, and Regional Hospital of Edea. ^12^ The registry collects clinical data including demographics, injury characteristics, prehospital care, injury severity, vital signs, and clinical outcomes. This data provides powerful insight into the landscape of prehospital care as it currently stands in Cameroon to drive evidence-based investment in health programs. Having now developed a robust trauma registry, we are uniquely positioned to study the clinical impact of LFR program implementation to determine its appropriateness as the first step toward formal trauma system development. In this study, we aim to characterize existing deficits in prehospital care in Cameroon to determine the appropriateness of LFR program implementation, determine a target LFR trainee population, and identify potential interventions to prioritize to inform curriculum development.

## METHODS

### Study setting

Cameroon is a Central African nation with a population over twenty-seven million and no established prehospital care system. Ambulance access is concentrated in its largest cities and use is generally limited to patient transport rather than emergency response. ^6^ The current practices for bringing injury victims to the hospital in the absence of formal prehospital infrastructure is unexplored in Cameroon.

### Cameroon Trauma Registry

We extracted data from a prospective multicenter cohort study of trauma patients (the Cameroon Trauma Registry (CTR)). At time of analysis in May 2023, data had been collected from four hospitals with diverse patient populations: Laquintinie Hospital, Regional Hospital of Limbe, Catholic Hospital of Pouma, and Regional Hospital of Edea. The database contains patients from January 2020 to December 2022. Inclusion criteria include patients of all ages presenting to the hospital for injuries who were admitted to the hospital, died in the emergency department, left against medical advice, or were transferred to another hospital. Patients were excluded if they were discharged directly from the emergency department. Patients enrolled in the registry following verbal consent were followed from hospital admission to discharge.

Data extracted from the CTR included demographics, injury characteristics including injury severity, clinical patterns, treatments, and outcomes. Patients who received prehospital care (PC) were compared to patients who did not receive prehospital care (NPC). Prehospital care was defined as informal medical intervention prior to presentation to a CTR center. The prehospital interventions assessed were bleeding control (including compression and elevation), fracture immobilization, tourniquet placement, C-spine immobilization, topical burn treatment, backboard support, CPR, assistance to the recovery position, and IV fluids. Data extracted from the CTR included demographics, injury characteristics including injury severity, clinical patterns, treatments, and outcomes. Patients who received prehospital care (PC) were compared to patients who did not receive prehospital care (NPC). Individuals who administered prehospital care were self-identified upon hospital presentation and included bystanders, individuals involved in the accident, family/friends, and those with informal medical training (police, military).

### Ethics statement

The University of California Los Angeles and Cameroonian Ministry of Public Health manage the CTR and ongoing ethical approval for the CTR is maintained at both institutions. Prior to enrollment in the CTR, local research assistants secured informed consents from patients or surrogates using an IRB-approved script. Research assistants encouraged patients and surrogates to ask questions. Participation in the CTR is completely voluntary and had no effect on medical care. Verbal consent was utilized given low risk for participation and variable literacy. Patients are excluded from CTR enrollment if they do not consent to participate. Patient information was deidentified at time of analysis.

### Data Analysis

Data were summarized using means and standard deviation for normally distributed numeric variables and by medians and interquartile ranges for nonparametric numeric variables. using the Shapiro-Wilk test. Frequencies and proportions were reported for categorical variables.

Comparisons between groups were made using Chi squared (no tail) for categorical variables and Kruskal-Wallis analysis for numerical variables. We evaluated associations between prehospital care and patient outcomes using logistic regression. For all analysis, alpha level of 0.05 was used for significance. Statistical analysis was conducted in Stata 17.

## RESULTS

In 2020, 2212 patients were enrolled in the CTR. The majority (1300, 76%) of injured patients were male and the median age was 31 (IQR 24,42). Overall, 88% (2033) lived in urban areas and 95% (2087) had family access to a cellphone. Road traffic injuries were most common injury mechanism (1580, 74%) and patients most often presented to the hospital setting with closed fractures (479, 22%), hematomas (340, 16%), and deep lacerations (334, 15%). The median travel distance to the hospital following injury was 10 km (IQR 5,13). (Table 1)

**Table 1.**
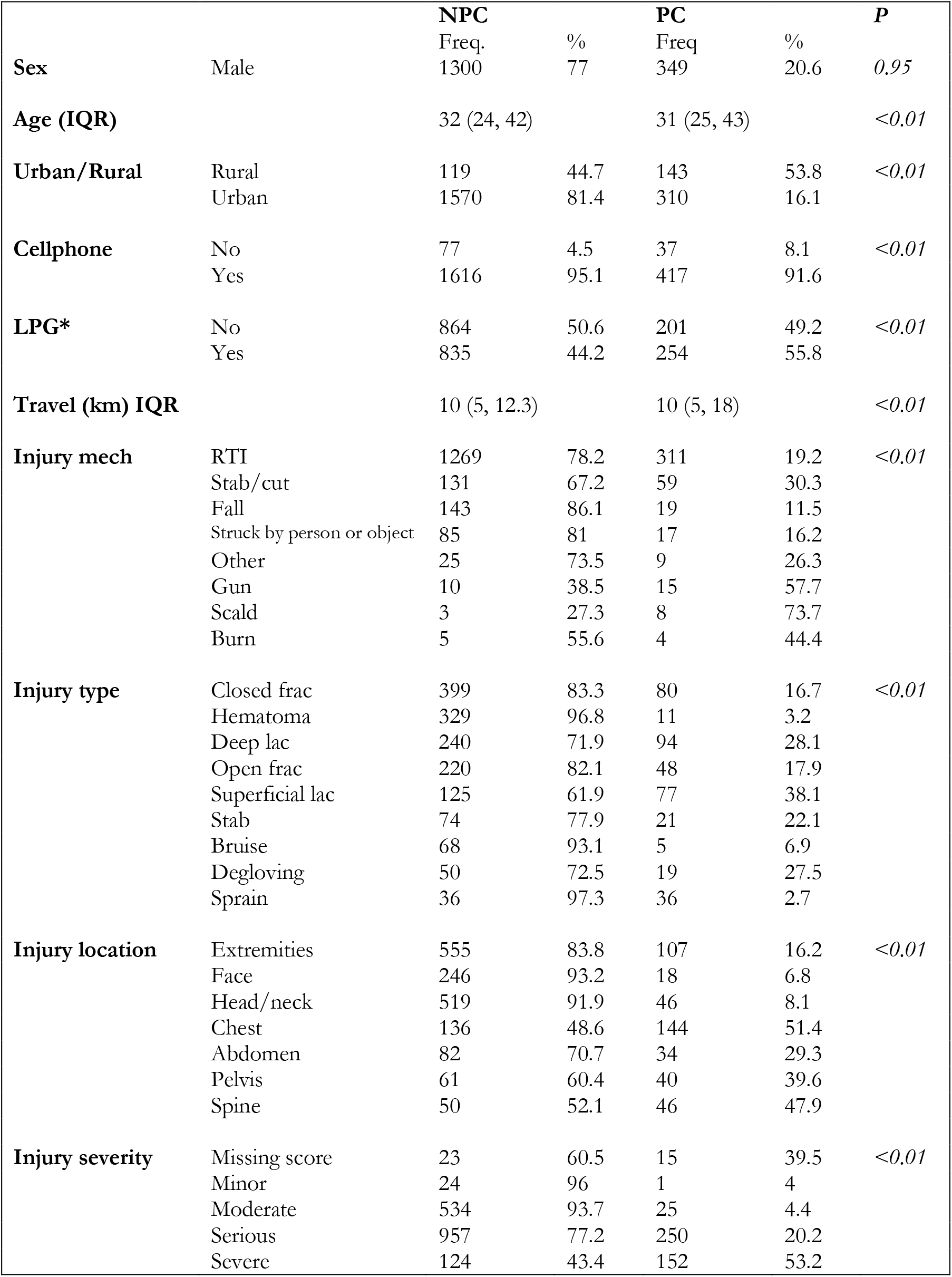
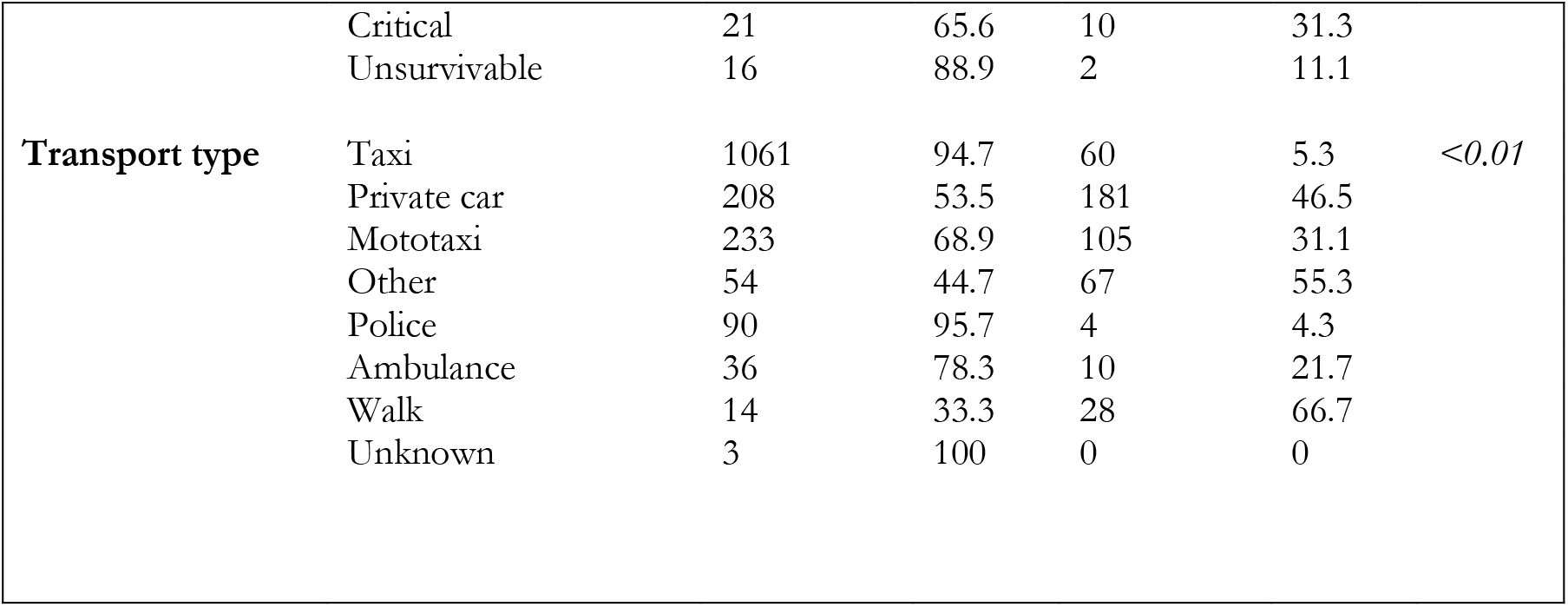
Demographics of trauma patients and prehospital care. * LPG = liquid petroleum gas, marker for socioeconomic status; all categorical variables reported as proportions and all numerical variables reported as medians with interquartile range

### Prehospital Care and Transport Patterns

Overall, 459 (20%) received and 1699 (76%) did not receive prehospital care. The vast majority (424, 93%) of prehospital care was provided by persons without formal training. Specifically, bystanders provided prehospital care for 70% (305), followed by relatives or friends (101, 23%) others involved in the accident (9, 2%), and those who self-identify as having medical training (9, 2%). The most common prehospital care interventions performed included bleeding control (370, 57%), fracture immobilization (139, 21%), and tourniquet placement (78, 12%). (Table 2)

**Table 2.**
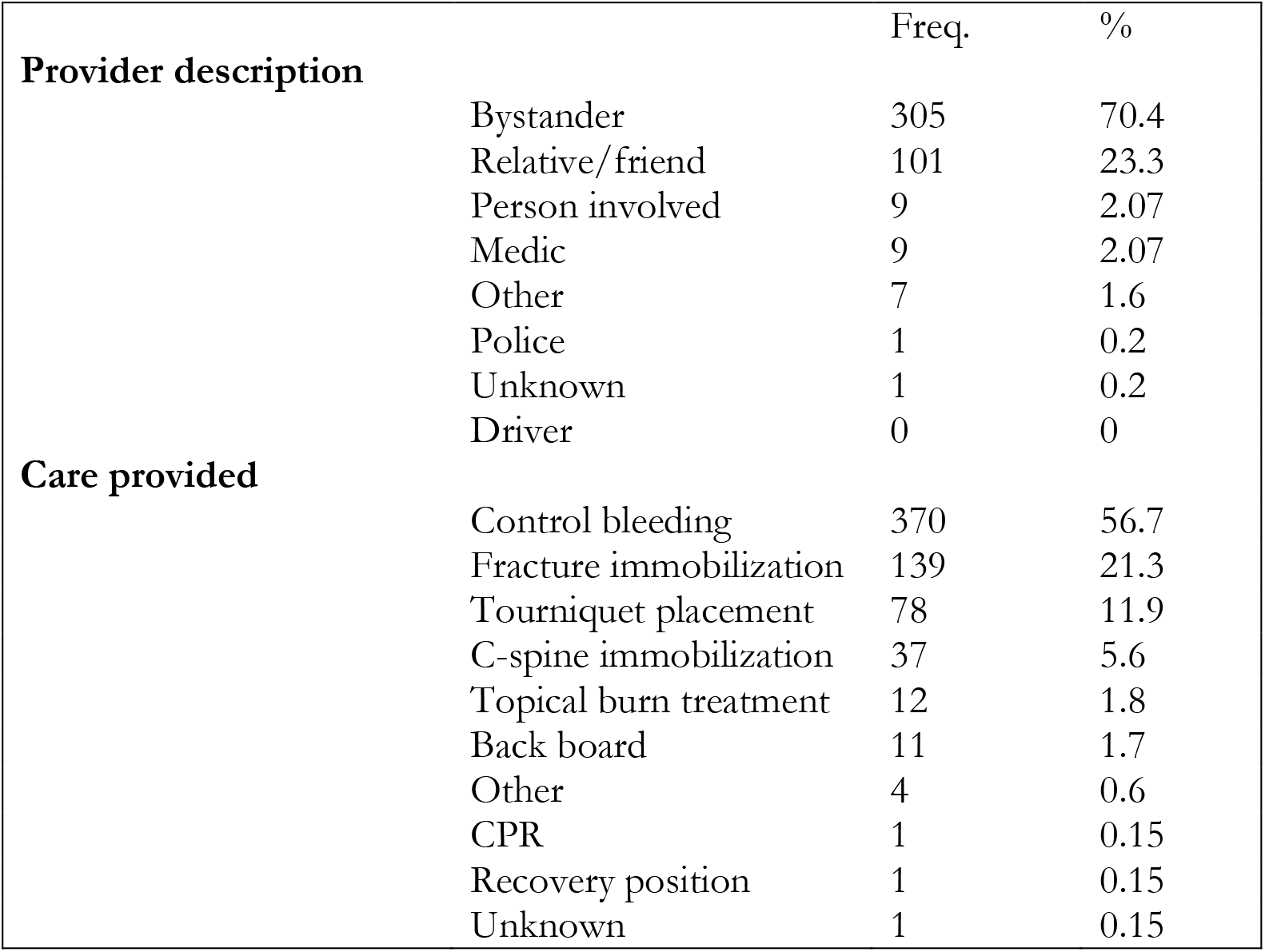
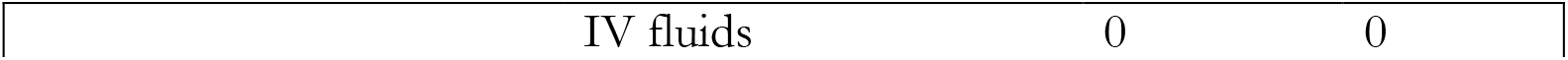
Classification of prehospital care providers and type of care provided.

| **Provider description** |  | Freq. | % |
| --- | --- | --- | --- |
|  | Bystander | 305 | 70.4 |
|  | Relative/friend | 101 | 23.3 |
|  | Person involved | 9 | 2.07 |
|  | Medic | 9 | 2.07 |
|  | Other | 7 | 1.6 |
|  | Police | 1 | 0.2 |
|  | Unknown | 1 | 0.2 |
|  | Driver | 0 | 0 |
| **Care provided** |  |  |  |
|  | Control bleeding | 370 | 56.7 |
|  | Fracture immobilization | 139 | 21.3 |
|  | Tourniquet placement | 78 | 11.9 |
|  | C-spine immobilization | 37 | 5.6 |
|  | Topical burn treatment | 12 | 1.8 |
|  | Back board | 11 | 1.7 |
|  | Other | 4 | 0.6 |
|  | CPR | 1 | 0.15 |
|  | Recovery position | 1 | 0.15 |
|  | Unknown | 1 | 0.15 |

|  |  |  |
| --- | --- | --- |
| IV fluids | 0 | 0 |

The most common means of transporting patients to the hospital was commercial vehicles including 52% by taxi (1152) and 15% by motorcycle taxi (339). Only 3% (57) were brought to the hospital by ambulance. (Table 1)

### Characteristics of prehospital care recipients

There were no significant differences in sex or age between PC and NPC. PC patients were more frequently from rural areas (54% vs. 45% p<0.0001) and did not have access to cellphones (91% vs. 95% p=0.003) but were users of Liquid Petroleum gas (55% vs. 44%, p= 0.003), a proxy of increased socioeconomic status. PC patients reported greater travel distance to the hospital NPC 15km15km±28.3 vs. PC 17km17km±34.9 p=0.0008). Prehospital care was significantly more common among patients transported by motorcycle taxi than by automobile taxis (31% vs. 5%, p<0.001). Just 20% of individuals transported by ambulance had prehospital care. (Table 1)

Additionally, PC more frequently presented with penetrating injury mechanisms (70% vs. 52% p<0.0001), including gunshot or stab wounds and both superficial and deep lacerations. PC recipients had increased severity of injury by highest estimated abbreviated injury score (PC 3.2±0.88 vs. NPC 2.7±0.80) (p<0.0001). (Table 1)

### Clinical consequences of prehospital care

Compared to NPC, the PC cohort presented with increased heart rate (NPC 87bpm±18.5 vs. PC 91bpm±14.5, p=0.0001) but reduced respiratory rate (NPC 23±10.3 vs. PC 22±4.4, p=0.0004) and systolic blood pressure (NPC 127mmHg±23.8 vs. PC 123mmHg±20.6, p=0.0001). Prehospital care recipients had increased rates of external bleeding (93% vs. 75%, p<0.0001), increased rates of abnormal breath sounds (4% vs. 1% p=0.001), and more abnormalities on primary survey overall (93% vs. 79%, p<0.0001). Despite this, the PC cohort had significantly increased rates of palpable pulse on presentation (99.8 vs. 98.6 p=0.042). With respect to disposition, PC recipients less frequently had a GCS<9 on primary survey (3% vs. 7%, p=0.003). (Table 3) Ultimately, multivariate logistic regression adjusted for injury severity identified PC cohort to be associated with increased injury survival. (OR 0.14, p<0.0001)

**Table 3.** Clinical consequences of prehospital care. * systolic blood pressure; ** heart rate; *** respiratory rate

|  |  | NPC |  | PC |  | P |
| --- | --- | --- | --- | --- | --- | --- |
|  |  | Mean |  | Mean |  |  |
| Vitals | SBP* | 128 | | 120 | | $<0.01$ |
| | HR** | 87 | | 91 | | $<0.01$ |
| | RR*** | 20 | | 22 | | $<0.01$ |
|  | Temp | 36.9 |  | 36.8 |  | 0.13 |
|  |  | Freq | % | Freq | % |  |
| Airway | Not patent | 31 | 2 | 4 | 1 | 0.16 |
|  | Yes patent | 1659 | 98 | 448 | 99 |  |
| Breathing | No chest rise | 17 | 1 | 1 | 0 | 0.27 |
|  | Abnormal chest rise | 1644 | 98 | 443 | 96 |  |
|  | Normal chest rise | 30 | 1 | 8 | 4 |  |
|  | Abnormal breath sounds | 28 | 98 | 19 | 96 | *<0.01* |
|  | Normal breath sounds | 1664 | 2 | 450 | 4 |  |
| **Circulation** | No palpable pulse | 23 | 1 | 1 | 0 | *0.04* |
|  | Yes palpable pulse | 1669 | 99 | 449 | 100 |  |
|  | No external bleeding | 408 | 24 | 30 | 7 | *<0.01* |
|  | Yes external bleeding | 1274 | 76 | 419 | 93 |  |
| **GCS** | ≤9 | 120 | 7.06 | 15 | 3 |  |
|  | >9 | 1579 | 93 | 440 | 97 |  |
|  |  |  |  |  |  | *<0.01* |
| **Overall** | No abnormalities | 363 | 21 | 34 | 7 |  |
|  | Yes abnormalities | 1336 | 79 | 421 | 93 |  |

## DISCUSSION

In this study we characterized existing patterns and gaps in prehospital care in Cameroon to support development of a tailored trainee LFR curriculum. Prehospital care is relatively uncommon and is currently provided mostly by untrained bystanders at the scene. The provision of prehospital care is higher among patient with severe injuries, with recipients of prehospital care less likely to die upon presentation than non-recipients. The efficacy of prehospital intervention, even in the absence of formal training, further justifies the need for investment and development of sustainable prehospital infrastructure in Cameroon.

To maximize access to prehospital care, LFR trainees must have high exposure to injuries as they arise. Considering that road traffic injuries constitute nearly three quarters of patients in our cohort, and commercial vehicles transport over half of injured patients, our findings suggest taxi and mototaxi drivers may be an ideal target trainee population in this setting. Mototaxi drivers have been trained as first responders elsewhere in sub-Saharan Africa, including Uganda, Chad, Nigeria, and Sierra Leone, with associated evidence of increase in access to prehospital care. Because the cultural landscape across sub-Saharan Africa varies so greatly and buy-in from commercial drivers is critical to the sustainability of the intervention, a 360-degree acceptability assessment is indicated to assess appropriateness of commercial drivers as LFR trainees in Cameroon.

The most common care provided by untrained responders was general bleeding control and those with active bleeding received more prehospital intervention, but tourniquet placement was far less common. Rapid hemorrhage control using tourniquets for compressible extremity hemorrhage in patients not yet in shock is strongly associated with reduced mortality. Hemorrhage management is an accessible and high-impact intervention that should be emphasized in the curricula for layperson first responder programs. ^13^ Using the same CTR dataset, our group previously demonstrated high rates of preventable deaths from hemorrhagic shock, which presents an addressable problem that informs curriculum development during future LFR trainings to reduce overall trauma mortality. In the absence of cardiac chain of survival infrastructure in Cameroon, CPR is low-yield for mortality benefits and will not be included in trainings. ^14^

Periodic review of clinical data from the CTR will remain critical to collaborating with local stakeholders for prehospital process planning. We plan on using the data to track rates of prehospital care, evaluate clinical outcomes, and analyze patterns of care among high-impact responders with future program implementation. The next step in our evaluation of the prehospital landscape will include qualitative analysis of the current system through stakeholder interviews to best assess the acceptability of potential prehospital interventions.

## LIMITATIONS

This study has several notable limitations. Due to the observational study design, we cannot infer causality in the relationship between prehospital care and clinical outcomes. Prehospital data collection for the trauma registry is completed by a registrar in the hospital, so some details about prehospital care in critically injured patients, including prehospital deaths, may be unknown.

## CONCLUSIONS

In Cameroon, prehospital care is provided by untrained bystanders to injury, but appears to generate improved clinical outcomes including reduced emergency department mortality. Commercial drivers provide most transport from the prehospital scene of injury to definitive care and have highest exposure to injury. Given the high rates of external bleeding among injury victims and the potential for impact with appropriate hemorrhage control, hemorrhage management will form a cornerstone of future LFR curricula. LFR program co-implementation in Cameroon with an existing trauma registry will permit ongoing quality improvement analyses for training not feasible in locations without access to a similar registry.

## Data Availability

We have attached all data, de-identified of patient information, from the Cameroon Trauma Registry, used in this analysis as a supplement to this manuscript

## ACKNOWLEDGEMENTS

Alain Chichom-Mefire and S. Ariane Christie are joint senior authors.

